# Genome-wide Polygenic Risk Score for Type 2 Diabetes in Indian Population

**DOI:** 10.1101/2023.02.24.23286351

**Authors:** Sandhya Kiran Pemmasani, Shravya Atmakuri, Anuradha Acharya

## Abstract

Genome-wide polygenic risk scores (PRS) for lifestyle disorders, like Type 2 Diabetes (T2D), are useful in identifying at-risk individuals early on in life, and to guide them towards healthier lifestyles. The current study was aimed at developing PRS for the Indian population using imputed genotype data from UK Biobank and testing the developed PRS on data from GenomegaDB™ of Indians living in India. 984 T2D cases and 2,818 controls were selected from Indian participants of UK Biobank to develop the PRS. Summary statistics available for South Asians, from the DIAMANTE consortium, were used to weigh genetic variants. LDpred2 algorithm was used to adjust the effect of linkage disequilibrium among the variants. The association of PRS with T2D, tested with logistic regression, was found to be very significant (AUC = 0.75, OR = 3.06 [95%CI = 2.78 - 3.36]). When participants were divided into four PRS quartile groups, the odds of developing T2D increased sequentially with higher PRS groups. The highest PRS group (top 25%) showed 6.6 fold increased risk compared to the rest of the participants (75%). The derived PRS was found to be significantly associated with T2D in the test dataset of 446 Indians (AUC = 0.6, OR = 1.48 [95%CI = 1.21 - 1.8]). Our results indicate the importance of developing population-specific PRS for T2D to stratify the individuals, and to recommend personalized preventive measures.

## INTRODUCTION

Type 2 diabetes (T2D) is one of the largest health emergencies in developing countries, and is considered as an avoidable pandemic of the 21st century [1,2]. According to the 2021 estimates of International Diabetes Federation (IDF), China and India have the highest numbers of people with diabetes [3]. It is further estimated that by 2045 the number of people with diabetes will have increased by 46%, with highest growth in middle-income countries. Economic development, urbanization and changed food habits could be the reasons for these increased numbers. In addition to that, genetics also plays a major role in increasing the prevalence of the disease. Several studies indicate that South Asians, in particular Asian Indians, are more susceptible to insulin resistance compared to other ethnic groups [2,4,5]. Even the migrant Indians living in different parts of the world were found to have higher diabetes rates [6,7].

Genome-wide association studies (GWAS) done so far on different populations have identified several single nucleotide polymorphisms (SNPs) associated with T2D. Odds ratios or effect sizes obtained from those studies are used to estimate the cumulative effect called polygenic risk score (PRS). It is a weighted sum of risk alleles and their estimated effect sizes [8]. Thus estimated PRS can be used to stratify the individuals into different risk groups, and to identify at-risk individuals. For the accurate estimation of PRS, effect sizes should be taken from GWAS done on the specific population under study. Due to lack of Indian-specific effect sizes, earlier research relied on European data. Recently, Mahajan et al [9] provided effect sizes, in terms of summary statistics, for different populations through DIAMANTE (DIAbetes Meta-ANalysis of Trans-Ethnic association studies) consortium. Their South Asian-specific summary statistics can be used to estimate PRS for the Indian population.

Before calculating a genome-wide PRS, effect sizes of SNPs are adjusted for linkage disequilibrium (LD) among them. LD is calculated by taking a reference dataset that is as close as possible to the population used to derive the summary statistics. Though large sample sizes are recommended for such a reference dataset, 1000 Genomes Phase 3 data with 489 South Asian individuals can be used for adjusting SNP effect sizes in the South Asian population. LDpred2 is a popular program to calculate genome-wide PRS using summary statistics and LD reference panel [10]. It uses a Bayesian algorithm to estimate posterior mean effect sizes from prior effect sizes of GWAS summary statistics. The ‘auto’ option of LDpred2 does not require any validation datasets to estimate the best-performing hyper-parameters. PRS thus calculated can further be utilized to build regression models to predict an individual’s genetic predisposition to the phenotype interest.

In this study, we have developed genome-wide PRS of T2D for the Indian population using UK Biobank data [11]. The developed PRS was tested on an independent dataset from GenomegaDB™ of Mapmygenome [12]. To our knowledge, this is the first study to systematically evaluate the utility of South Asian-specific summary statistics of T2D on the Indian population. The developed PRS can be used as a prognostic metric to identify high risk individuals early on in life, and to recommend personalized preventive measures.

## METHODOLOGY

### Study participants

#### UK Biobank

UK Biobank is a large, population-based prospective study, with over 500,000 participants, aged 40-69 years when recruited in 2006-2010, living in the United Kingdom. Extensive phenotypic and genotypic data of the participants was collected across four assessment visits. Data of 3,983 Indian participants (Field ID#: 20115) were used in the present study to build polygenic risk scores for T2D. Participants were excluded based on - mismatch between reported sex and genetic sex; sex chromosome aneuploidy; excessive or low heterozygosity; outliers based on 3 standard deviations from the mean of top 3 principal components; and relatedness with kinship coefficient > 0.088 [13]. Diabetic cases were identified based on International Classification of Diseases (ICD) codes 9 and 10, self-report, doctor diagnosis, HbA1C levels and medication for diabetes. Data fields and codes are given in Table 1 [14 -16]

**Table 1:** Selection of T2D cases from UK Biobank

| Field Name | Field ID | Code |
| --- | --- | --- |
| Diabetes diagnosed by a doctor | 2443 | 1 – Yes<br>0 – No<br>-1/-3/NA - Missing |
| Self-reported | 20002 | 1220, 1222, 1223,1276, 1468, 1607 |
| HbA1c | 30750 | >=48 mmol/mol<br>NA - Missing |
| Medication for cholesterol, blood pressure, diabetes, or take exogenous hormones [Female Question] | 6153 | 3<br>-1/-3/NA - Missing |
| Medication for cholesterol, blood pressure, diabetes [Male Question] | 6177 | 3<br>-1/-3/NA - Missing |
| Treatment/Medication | 20003 | 1140857494,1140857496,1140857500,1140857502,1140857506,1140857584,1140857586,1140857590,1140874646,1140874650,1140874652,1140874658,1140874660,1140874664,1140874666,1140874674,1140874678,1140874680,1140874686,1140874690,1140874706,1140874712,1140874716,1140874718,1140874724,1140874726,1140874728,1140874732,1140874736,1140874740,1140874744,1140874746,1140883066,1140884600,1140921964,1141152590,1141153254,1141153262,1141156984,1141157284,1141168660,1141168668,1141169504,1141171508,1141171646,1141171652,1141173786,1141173882,1141177600,1141177606,1141189090,1141189094 |
| ICD10 | 41270 | E10 – E14 |
| ICD9 | 41271 | 250 |

#### GenomegaDB™

GenomegaDB™ of Mapmygenome is a genotype and phenotype database of Indians living in India. Genotype data was generated using Illumina’s HumanCoreExome-12 (HCE-12), HumanCoreExome-24 (HCE-24) and Infinium Global Screening Array-24 (GSA-24). Phenotype data was collected through a printed questionnaire that included individual clinical history, operative procedures, medications, family history, country of birth, among others. Written informed consent, including the consent to use data for research, was taken from each individual. In the current study, samples processed on GSA-24 arrays version 1.0, 2.0 and 3.0 were considered. Standard QC on samples included - removing samples with low call rate (<95%), gender mismatch, extreme heterozygosity, relatedness or that were outliers in principal component analysis (PCA). Diabetic cases and controls, aged more than 30 years, were selected based on self-reported clinical history and medications.

### Genotype data

#### UK Biobank

UK Biobank v3 imputed data, available in BGEN v1.2 format, was used in the analysis (Field ID#: 22828). Only the variants that overlap with the ones present on GSA chips were considered. QCTOOL v2 [17] was used to retrieve the samples and variants of interest. Further filtration was done for INFO score >=0.3 and minor allele frequency >= 0.05 (Figure 1)

**Figure 1:**
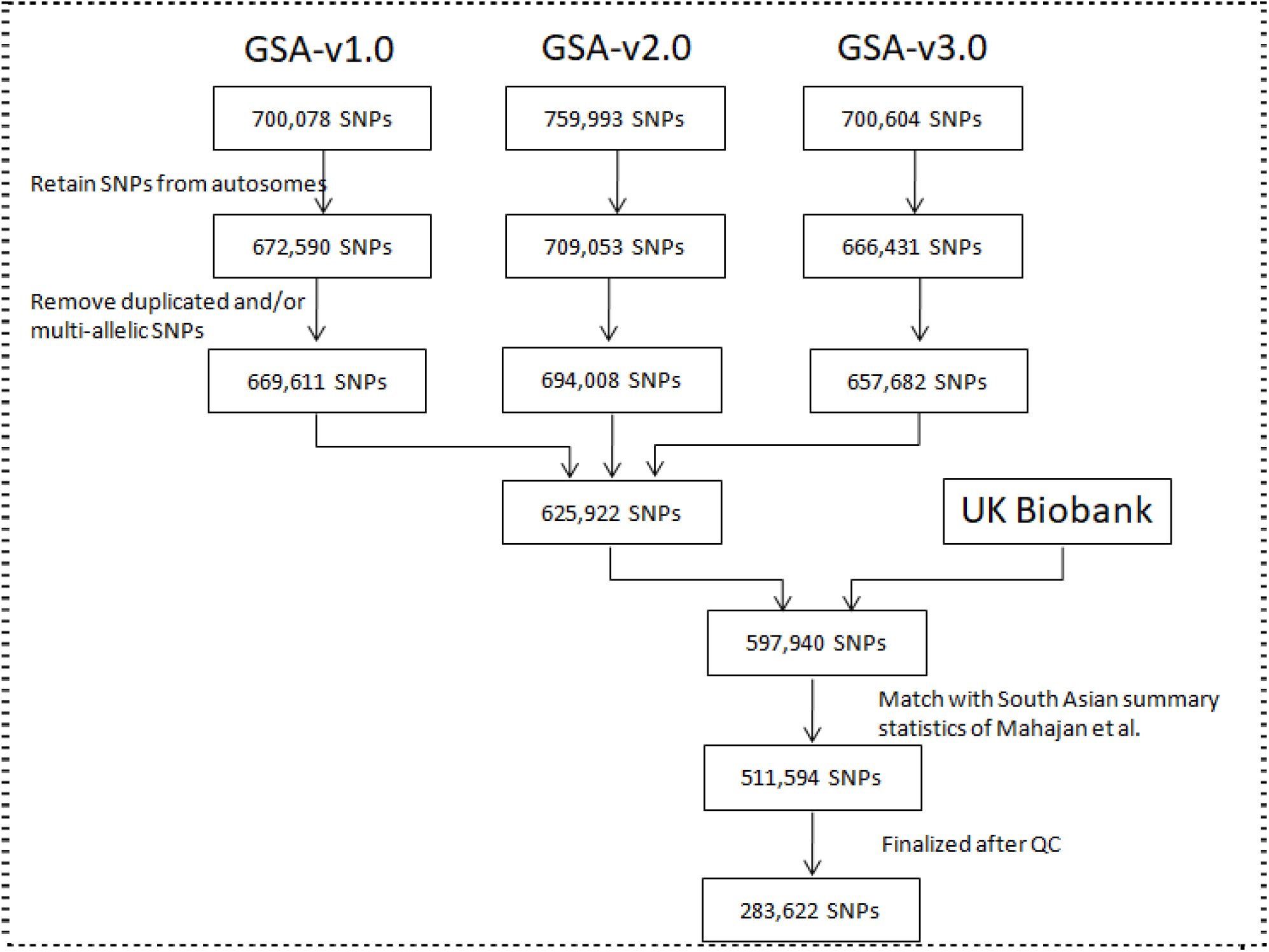
SNP Selection

#### GenomegaDB™

625,962 autosomal bi-allelic SNPs that were genotyped across the three versions of GSA chip were considered in the analysis. Genotypes were phased using SHAPEIT v2.15 [18], and missing ones were imputed with IMPUTE v2.3 software [19], using 1000 Genomes Phase 3 data of South Asians as reference (Figure 1)

### Summary statistics

Summary statistics from South Asian-specific GWAS meta-analysis released by Mahajan et al, with 16,540 cases and 32,952 controls, were obtained from DIAMANTE consortium [9]. QC on summary statistics was done as per the method proposed by Prive et al 2022 [20]. Effective sample size (neff) was calculated as 4 / ((1 / cases) + (1 / controls)). Then, standard deviation of genotypes (sd_ss) was calculated as 2 / sqrt(n_eff_ * beta_se^2 + beta^2)), where ‘beta’ is the effect size and ‘beta_se’ is the standard deviation of effect size. Standard deviation from the allele frequencies (sd_af) was calculated as sqrt(2*f*(1-f)). Variants were filtered out if sd_ss < (0.7 * sd_af) or sd_ss > (sd_af + 0.1) or sd_ss < 0.1 or sd_af < 0.05. Variants were also filtered out if the absolute difference in allele frequencies of the UK Biobank data and the frequencies given in summary statistics was > 0.1.

### LD Reference panel

1000 Genomes Phase 3 data in PLINK format was obtained through PLINK2 resources [21]. To increase the predictive power of PRS, the South Asian panel (SAS), composed of 489 individuals, was considered.

### Polygenic Risk Scores (PRS)

Polygenic risk scores were generated using the LDpred2 algorithm implemented in ‘bigsnpr’ package (version 1.11.6) of R [22,23]. The ‘*auto*’ option, which directly estimates the model parameters from the data without the requirement of training data, was used along with *shrink_corr = 0*.*95 and allow_jump_sign = FALSE*, as per the procedure recommended by LDpred2 authors [24]. PRS were normalized to have mean zero and standard deviation one.

### Prediction of Type 2 Diabetes

Univariate logistic regression model was built to understand the association of PRS with T2D. Model accuracy was assessed using standard receiver operating curves (ROC). Analyses were done with R v4.2.

## RESULTS

Out of 4,161 Indian participants of UK Biobank, genotype data was available for 3,983 participants. After the sample QC, 3,802 samples were included in the final analysis, of whom 984 were T2D cases and 2,818 were controls. 598,319 autosomal SNPs, with INFO score >=0.3, and overlapping with SNPs of Illumina’s GSA arrays versions 1.0, 2.0 and 3.0, were considered in the analysis. South Asian specific GWAS summary statistics obtained from the DIAMANTE consortium contains information on 10,401,621 SNPs. QC on summary statistics and UK Biobank genotype data resulted in 283,622 SNPs, which were finally used in developing genome-wide PRS (Figure 1).

The LDpred2 algorithm, along with the South Asian 1000 Genomes LD Reference panel, was used to correct the effect sizes given in summary statistics. PRS for each sample was calculated as a sum of the number of risk alleles weighted by the adjusted effect sizes. Figure 2A shows the distribution of normalized PRS in cases and controls.

**Figure 2:**
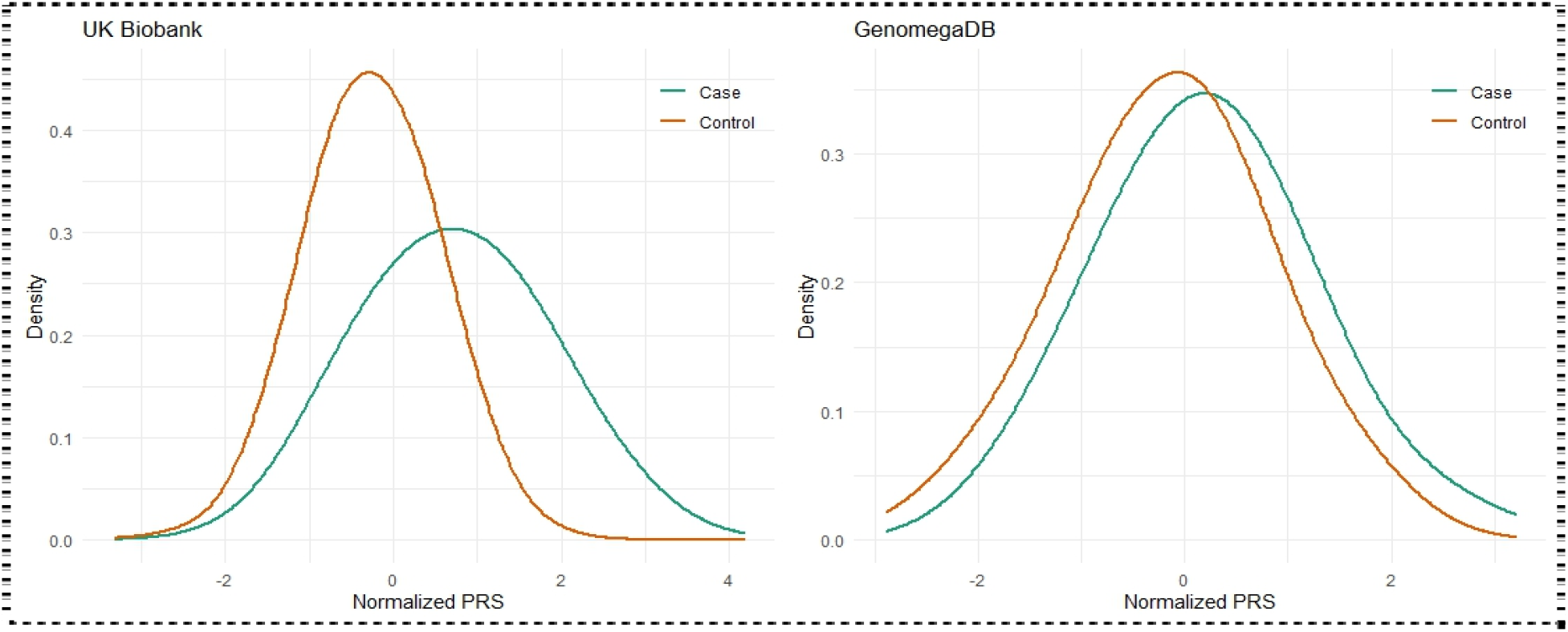
Distribution of normalized PRS. A) UK Biobank B) GenomegaDB™

The association analysis of PRS with T2D resulted in OR/SD of 3.06 (95% CI: 2.83 - 3.32) and AUC of 0.753 (Table 2 and Figure 3). When samples were divided into PRS quartiles, and the lowest quartile was taken as reference, all sequential PRS groups showed high risk of developing T2D (Table 3). The risk of developing T2D was 9.32 fold higher in the participants of the fourth quartile (top 25%) when compared with the participants of the first quartile (bottom 25%). The risk was 6.62 fold higher when the top 25% of participants were compared with the rest of 75%.

**Table 2:** Association analysis of genome-wide PRS with Type 2 Diabetes

| Dataset | Beta | SE (Beta) | OR/SD | AUC |
| --- | --- | --- | --- | --- |
| UK Biobank | 1.12 | 0.04 | 3.06 | 0.75 |
| GenomegaDB™ | 0.39 | 0.1 | 1.48 | 0.6 |

**Table 3:** Association analysis across different quartiles of PRS

| Reference Quartile | High PRS Quartile | UK Biobank |  |  | GenomagaDB™ |  |  |
| --- | --- | --- | --- | --- | --- | --- | --- |
|  |  | OR/SD | 95% CI | P-Value | OR/SD | 95% CI | P-Value |
| 1st quartile | 2nd quartile | 1.28 | 0.98 - 1.67 | 0.0688 | 1.84 | 1.07 - 3.17 | 0.0286 |
| 1st quartile | 3rd quartile | 2.00 | 1.56 - 2.58 | 5.49e-08 | 1.56 | 0.9 - 2.7 | 0.1145 |
| 1st quartile | 4th quartile | 9.32 | 7.37 - 11.78 | < 2e-16 | 2.78 | 1.6 - 4.81 | 0.0002 |
| 1st, 2nd, 3rd quartiles (Bottom 75%) | 4th quartile (Top 25%) | 6.62 | 5.63 - 7.79 | < 2e-16 | 1.95 | 1.26 - 3 | 0.0026 |

**Figure 3:**
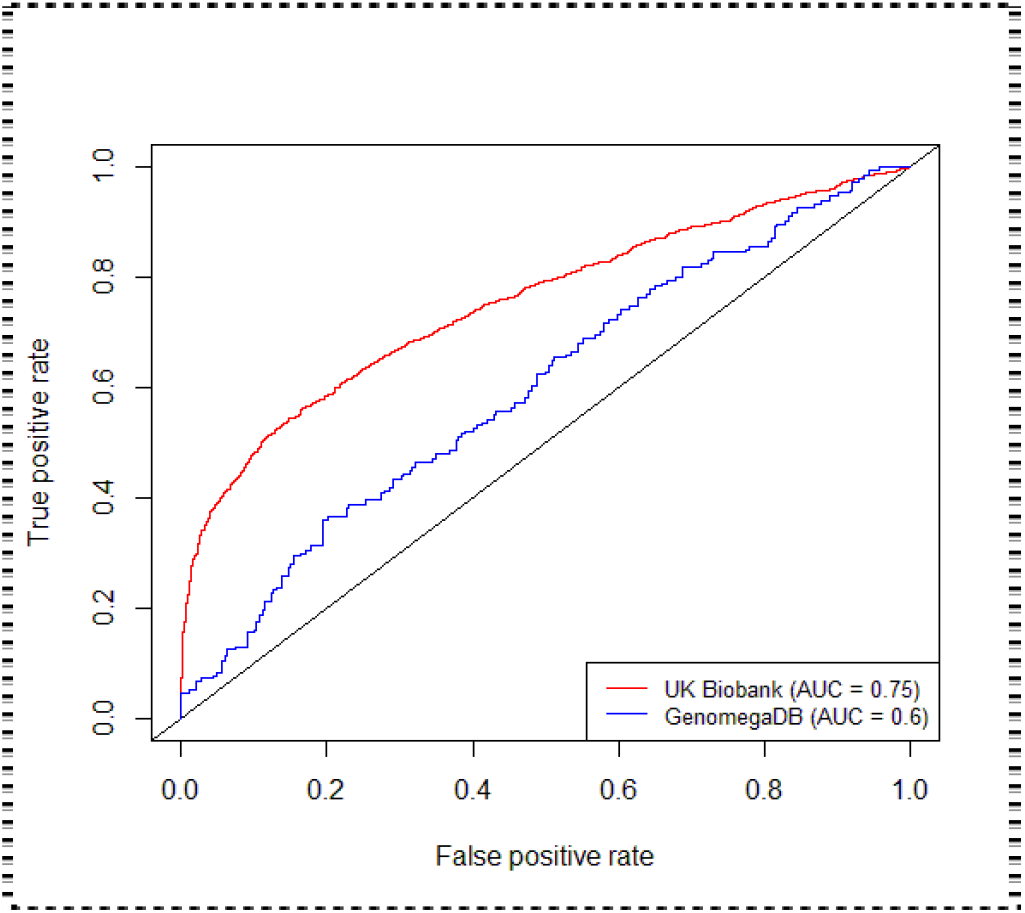
AUCs of PRS developed on data from UK Biobank and GenomegaDB™

In order to test the performance of PRS in an independent dataset, 327 cases and 396 controls were selected from GenomegaDB™ of Mapmygenome. After sample QC, we were left with 194 cases and 252 controls. Figure 2B shows the distribution of normalized PRS in cases and controls of GenomegaDB™. Association analysis showed OR/SD of 1.48 ((95% CI: 1.21 - 1.8). The risk of developing T2D was 2.78 fold higher in samples of the fourth quartile (top 25%) when compared with samples of the first quartile (bottom 25%).

## Data Availability

This research has been conducted using the UK Biobank Resource under Application Number 81481

## DISCUSSION

In this study, we derived genome-wide PRS of T2D for the Indian population, using case-control samples available at UK Biobank. LDpred2 algorithm, with weights extracted from South Asian summary statistics of DIAMENTE consortium, gave PRS that was significantly associated with T2D (AUC: 0.75). Participants in the fourth PRS quartile (top 25%) showed 6.62 fold increase in genetic risk compared to the rest of 75%. Data from GenomegaDB™ was used to validate the PRS, and to replicate the association. In spite of smaller sample size, it supported the hypothesis that PRS plays a major role in predicting the onset of diabetes. It showed 1.95 fold increased risk of diabetes in the top quartile (top 25%) compared to the rest of 75%. This indicates the importance of PRS in stratifying the individuals into different risk groups.

Assessment of UK Biobank participants was done at four different time points. Availability of follow-up data, along with data from different questionnaires and biochemical assays, allowed the reliable detection of diabetic patients. In the case of GenomegaDB^M^, assessment of T2D status was purely based on self-report, which might result in misclassification. In spite of lacking follow-up data, GenomegaDB™ has the advantage of coming from Indians living in India. For lifestyle disorders, like diabetes, it is preferable to take data from the native population having the same lifestyle and environment to that of the population for which inferences are made.

The biggest hurdle in developing genome-wide PRS for the Indian population is lack of summary statistics for SNP associations with T2D. Predictive ability of PRS is compromised if the effect sizes and frequencies are taken from other population groups. Earlier study done by Lamri et al [24] on prediction of gestational diabetes in South Asian women showed that the accuracy of PRS was higher with mutli-ethnic summary statistics that includes South Asian samples than that of European. Similar results were observed by Hodgson et al [25] while constructing T2D PRS for British Pakistanis and Bangladeshis. Now, the availability of South Asian summary statistics from the DIAMENTE consortium facilitated the development of a framework for accurate estimation of PRS for the Indian population. Further large scale studies are warranted to estimate SNP effect sizes from the Indians living in India.

LDpred2-auto method makes the construction of PRS an easy process compared to its counter-methods LDpred2-inf and LDpred2-grid which need validation data to estimate the hyper-parameters. The recent publication from Prive et al [20] gave many suggestions on improving the performance of the algorithm. Especially, quality control on summary statistics improves the predictive performance of PRS. Though LD metrics calculated from South Asian samples of 1000 Genomes project were used in this study, a bigger dataset is recommended.

In conclusion, irrespective of data from UK Biobank or GenomegaDB™, our PRS showed high accuracy in predicting the risk of developing T2D. Results indicated that genome-wide PRS holds strong potential to be adopted in clinical care to identify high risk individuals and in early intervention to guide towards healthier lifestyles.

## REFERENCES

1. Pradeepa R, Mohan V. Epidemiology of type 2 diabetes in India. Indian J Ophthalmol. 2021 Nov; 69(11):2932–2938. doi: 10.4103/ijo.IJO_1627_21. PMID: 34708726; PMCID: PMC8725109.

2. Unnikrishnan R, Pradeepa R, Joshi SR, Mohan V. Type 2 Diabetes: Demystifying the Global Epidemic. Diabetes. 2017;66(6):1432–1442. doi:10.2337/db16-0766

3. Sun H, Saeedi P, Karuranga S, et al. IDF Diabetes Atlas: Global, regional and country-level diabetes prevalence estimates for 2021 and projections for 2045. Diabetes Res Clin Pract. 2022;183:109119. doi:10.1016/j.diabres.2021.109119

4. Joseph A, Thirupathamma M, Mathews E, Alagu M. Genetics of type 2 diabetes mellitus in Indian and Global Population: A Review. The Egyptian Journal of Medical Human Genetics. 2022;23(1):135. doi:10.1186/s43042-022-00346-1

5. Wells JC, Pomeroy E, Walimbe SR, Popkin BM, Yajnik CS. The Elevated Susceptibility to Diabetes in India: An Evolutionary Perspective. Front Public Health. 2016;4:145. Published 2016 Jul 7. doi:10.3389/fpubh.2016.00145

6. Mohan V. Why are Indians more prone to diabetes?. J Assoc Physicians India. 2004;52:468–474.

7. Abate N, Chandalia M. Ethnicity, type 2 diabetes & migrant Asian Indians. Indian J Med Res. 2007;125(3):251–258.

8. Zhang C, Ye Y, Zhao H. Comparison of Methods Utilizing Sex-Specific PRSs Derived From GWAS Summary Statistics. Front Genet. 2022;13:892950. Published 2022 Jul 8. doi:10.3389/fgene.2022.892950

9. Mahajan A, Spracklen CN, Zhang W, et al. Multi-ancestry genetic study of type 2 diabetes highlights the power of diverse populations for discovery and translation. Nat Genet. 2022;54(5):560–572. doi:10.1038/s41588-022-01058-3

10. Privé F, Arbel J, Vilhjálmsson BJ. LDpred2: better, faster, stronger [published online ahead of print, 2020 Dec 16]. Bioinformatics. 2020;36(22-23):5424–5431. doi:10.1093/bioinformatics/btaa1029

11. Sudlow C, Gallacher J, Allen N, et al. UK biobank: an open access resource for identifying the causes of a wide range of complex diseases of middle and old age. PLoS Med. 2015;12(3):e1001779. Published 2015 Mar 31. doi:10.1371/journal.pmed.1001779

12. https://mapmygenome.in/

13. Purcell S, Neale B, Todd-Brown K, et al. PLINK: a tool set for whole-genome association and population-based linkage analyses. Am J Hum Genet. 2007;81(3):559–575. doi:10.1086/519795

14. Tamlander M, Mars N, Pirinen M; FinnGen Widén E, Ripatti S. Integration of questionnaire-based risk factors improves polygenic risk scores for human coronary heart disease and type 2 diabetes. Commun Biol. 2022;5(1):158. Published 2022 Feb 23. doi:10.1038/s42003-021-02996-0

15. Eastwood SV, Mathur R, Atkinson M, et al. Algorithms for the Capture and Adjudication of Prevalent and Incident Diabetes in UK Biobank. PLoS One. 2016;11(9):e0162388. Published 2016 Sep 15. doi:10.1371/journal.pone.0162388

16. Peakman TC, Elliott P. The UK Biobank sample handling and storage validation studies. Int J Epidemiol. 2008;37 Suppl 1:i2–i6. doi:10.1093/ije/dyn019

17. https://www.well.ox.ac.uk/~gav/qctool_v2/

18. O. Delaneau, J. Marchini, JF. Zagury (2012) A linear complexity phasing method for thousands of genomes. Nat Methods. 9(2):179–81. doi: 10.1038/nmeth.1785

19. B. Howie, C. Fuchsberger, M. Stephens, J. Marchini, and G. R. Abecasis (2012) Fast and accurate genotype imputation in genome-wide association studies through pre-phasing. Nature Genetics 44(8): 955–959

20. Privé F, Arbel J, Aschard H, Vilhjálmsson BJ. Identifying and correcting for misspecifications in GWAS summary statistics and polygenic scores. HGG Adv. 2022;3(4):100136. Published 2022 Aug 18. doi:10.1016/j.xhgg.2022.100136

21. https://cran.r-project.org/web/packages/plinkQC/vignettes/Genomes1000.pdf

22. Privé F, Aschard H, Ziyatdinov A, Blum MGB. Efficient analysis of large-scale genome-wide data with two R packages: bigstatsr and bigsnpr. Bioinformatics. 2018;34(16):2781–2787. doi:10.1093/bioinformatics/bty185

23. R Core Team (2021). R: A language and environment for statistical computing. R Foundation for Statistical Computing, Vienna, Austria.

24. https://privefl.github.io/bigsnpr/articles/LDpred2.html

25. Hodgson S, Huang QQ, Sallah N, et al. Integrating polygenic risk scores in the prediction of type 2 diabetes risk and subtypes in British Pakistanis and Bangladeshis: A population-based cohort study. PLoS Med. 2022;19(5):e1003981. Published 2022 May 19. doi:10.1371/journal.pmed.1003981

